# Modelling the population-level protection conferred by COVID-19 vaccination

**DOI:** 10.1101/2021.03.16.21253742

**Authors:** Pranesh Padmanabhan, Rajat Desikan, Narendra M. Dixit

## Abstract

Although severe acute respiratory syndrome coronavirus 2 (SARS-CoV-2) vaccines work predominantly by eliciting neutralizing antibodies (NAbs), how the protection they confer depends on the NAb response to vaccination is unclear. Here, we collated and analysed *in vitro* dose-response curves of >70 NAbs and constructed a landscape defining the spectrum of neutralization efficiencies of NAbs elicited. We mimicked responses of individuals by sampling NAb subsets of known sizes from the landscape and found that they recapitulated responses of convalescent patients. Combining individual responses with a mathematical model of within-host SARS-CoV-2 infection post-vaccination, we predicted how the population-level protection conferred would increase with the NAb response to vaccination. Our predictions captured the outcomes of vaccination trials. Our formalism may help optimize vaccination protocols, given limited vaccine availability.

**One sentence summary:** Viremic control by the spectrum of neutralizing antibodies elicited by vaccination determines COVID-19 vaccine efficacies.

---

Approved SARS-CoV-2 vaccines have shown remarkable but varying efficacies in clinical trials, reducing the incidence of symptomatic infections by 62-96% (*1-4*). The protection has been found to be predominantly due to NAbs elicited by the vaccines; cellular immunity appeared to play a secondary role (*1, 2*). The NAb response elicited by primary SARS-CoV-2 infection is diverse, spanning >1000-fold variation in Ab titres and *in vitro* neutralization efficiencies across individuals (*5, 6*), and appears not to correlate with disease severity (*7*). NAb titres following vaccination were comparable to or even lower at times than those from convalescent patients (*1, 2, 8*). The protection accorded by the vaccines is thus surprising. It is possible, based on animal studies (*9*), that lower NAb titres are protective at the time of challenge than post infection. Knowledge of how the level of protection depends on the NAb titres and their neutralization efficiencies is lacking. This knowledge gap hinders rational optimization of vaccination protocols, which is important today given limited vaccine supplies (*10*). Here, we developed a mathematical model that quantitatively predicts the population-level protection conferred by vaccines as a function of the NAb responses they elicit.

A major challenge to describing the effects of vaccination is the diversity of the NAb responses elicited; no formalism exists to predict the diversity or its effects on protection. We addressed this challenge by adapting the classic idea of *shape space*, which has aided quantification of the immune repertoire (*11*), for characterizing NAbs. Accordingly, we sought features, also termed shape parameters, of the NAbs that would predict their neutralization efficiencies. Numerous studies have isolated individual NAbs from patients and assessed their neutralization efficiencies *in vitro*, with the aim of developing NAbs for therapeutic applications. We compiled dose-response curves (DRCs) of >70 NAbs thus isolated and fit them using the standard sigmoidal function as well as the median-effect equation (*12*) (materials and methods, fig. S1, table S1). The equations fit the data well (Fig. 1A, and figs. S2 and S3), indicating that two parameters, the 50% inhibitory concentration, *IC*_50_, and the slope, *m*, of the DRC, were sufficient to characterize the neutralization efficiency of the NAbs (Fig. 1A and table S1). The best-fit *IC*_50_ and *m* varied widely across NAbs (Fig. 1B). *IC*_50_ ranged from ∼10^−3^ µg/ml to ∼140 µg/ml (Fig. 1B), in close agreement with reported estimates, giving us confidence in the fits (fig. S4A and table S1). *m*, the importance of which has been recognized with HIV-1 and hepatitis C (*12, 13*) but has not typically been reported for SARS-CoV-2, spanned the range of ∼0.2 to 2 (Fig. 1). This variability in *IC*_50_ and *m* was not restricted to a particular pseudotyped virus construct or backbone used (fig. S4, B and C), the cell line used (fig. S4, D and E), or assay conditions, which could vary across studies (fig. S4, F and G). The variability was thus intrinsic to the NAbs, indicating the spectrum of NAbs elicited. Furthermore, akin to HIV-1 antibodies (*12*), the variations in *IC*_*50*_ and *m* of the SARS-CoV-2 NAbs appeared independent. For instance, the NAbs BD-361 and REGN10954 had similar *IC*_50_ (both ∼0.04 µg/ml), but vastly different *m* (∼0.7 and ∼1.5, respectively), whereas the NAbs CC12.3 and 515-5 had vastly different *IC*_50_ (∼0.02 µg/ml and 1.6 µg/ml, respectively), but similar *m* (both ∼1). *IC*_*50*_ and *m* were thus not only sufficient but also necessary for quantifying the neutralization efficiencies of NAbs. We therefore employed *IC*_*50*_ and *m* as the required shape parameters. Plotting the NAbs on an *IC*_50_-*m* plot, we identified the NAb shape space (Fig. 2), which, because of its two-dimensional nature, we termed the ‘landscape of SARS-CoV-2 NAbs’.

**Figure 1.**
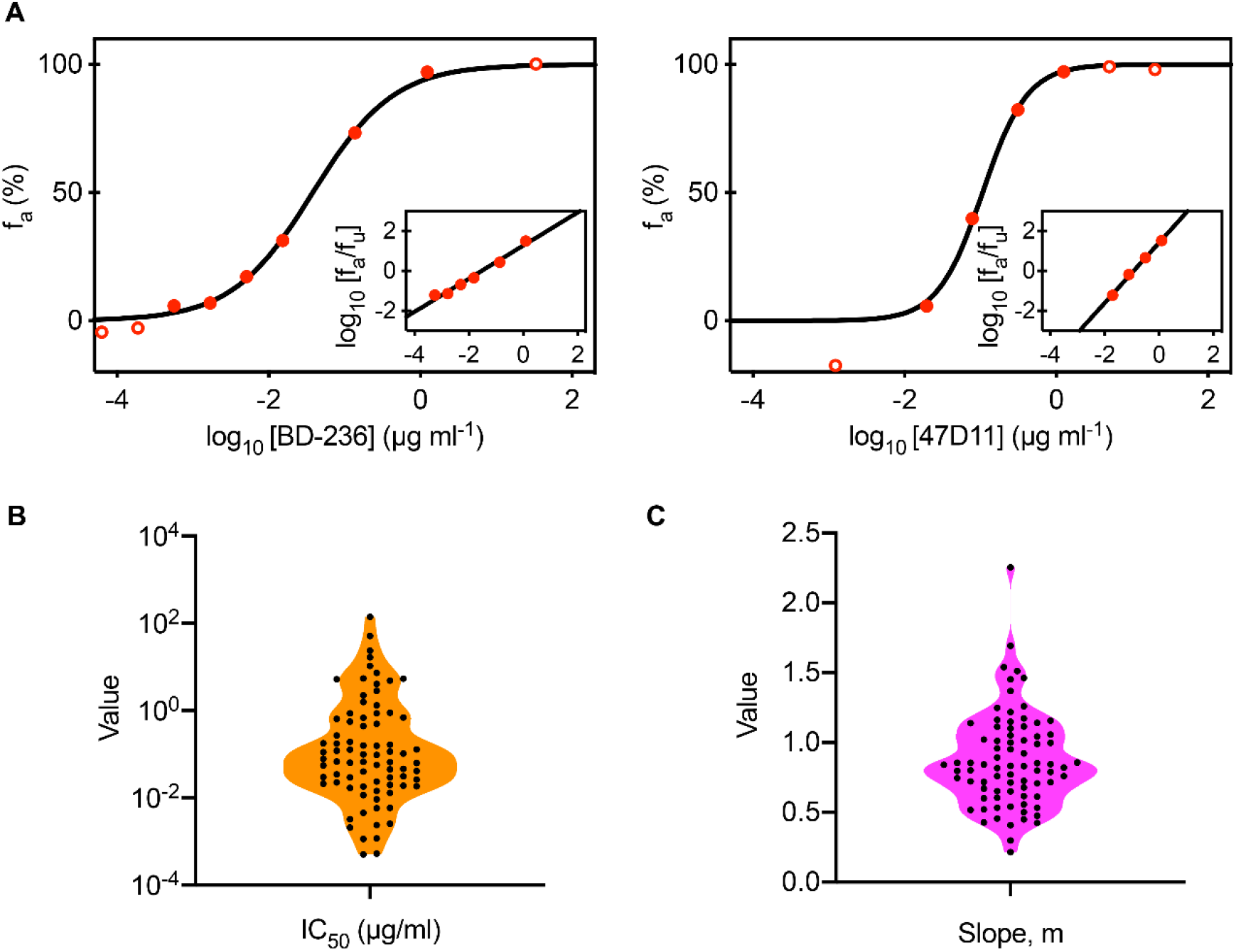
Analysis of dose-response curves of SARS-CoV-2 NAbs. (A) Fits (lines) of the standard sigmoidal equation and the median-effect equation (*inset*) to published experimental data (circles) of the fraction of infection events blocked, *f*_*u*_, as a function of NAb concentration, shown for two NAbs, BD-236 (left) and 47D11 (right). Experimental data points with 1% < *f*_*u*_ < 99% (filled circles) were considered for parameter estimation. Fits for the remaining NAbs are in figs. S2 and S3. The best-fit estimates of (B) *IC*_50_ and (C) *m* for all the NAbs analysed.

**Figure 2.**
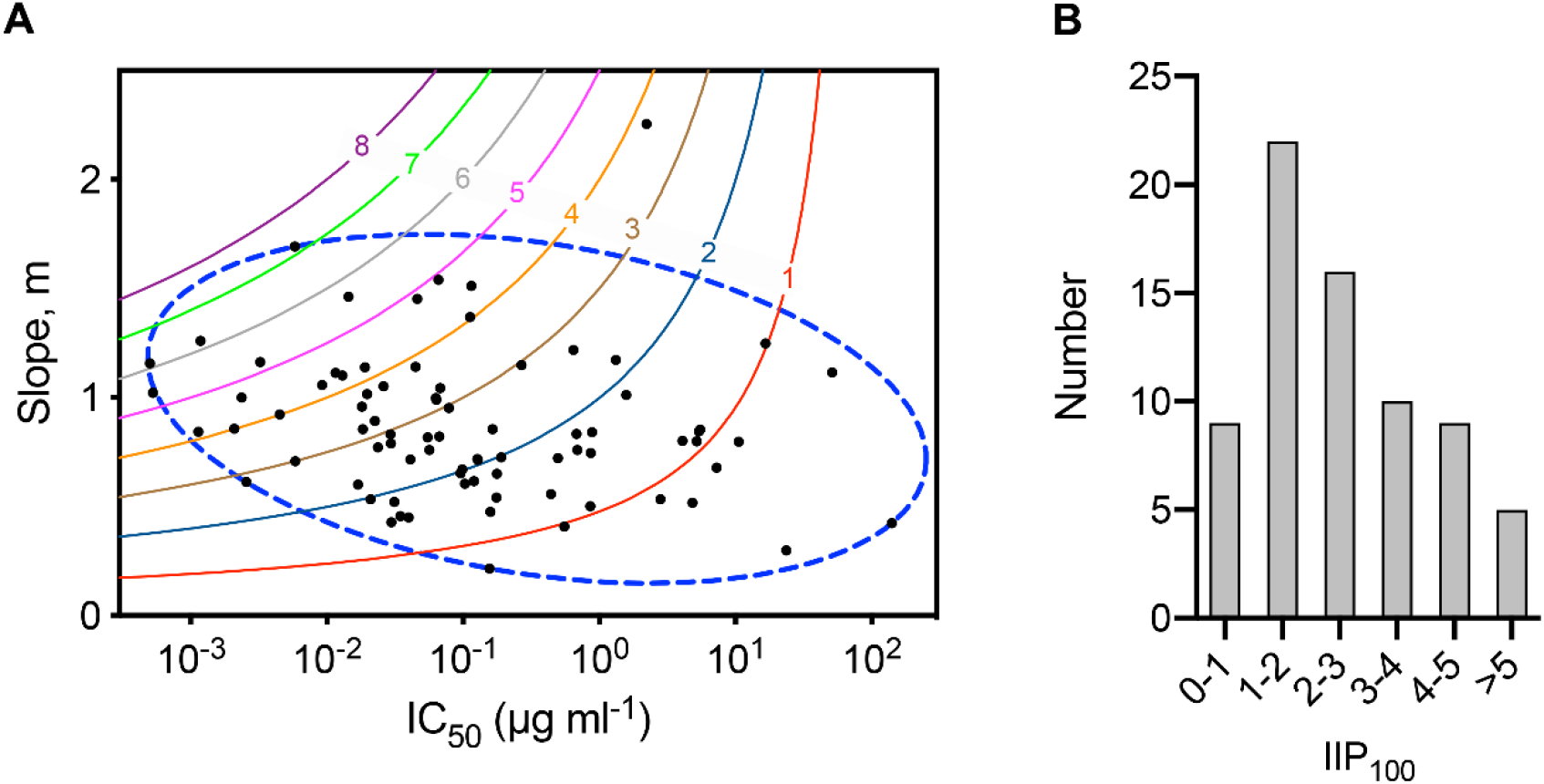
The landscape of SARS-CoV-2 NAbs. (A) SARS-CoV-2 NAbs analysed in Fig. 1 depicted on an *IC*_50_-*m* plot. Each dot represents a NAb. 8 NAbs that have multiple neutralisation curves reported are represented multiple times (table S1). Solid lines are loci of points corresponding to fixed *IIP* values computed at 100 µg/ml. The ellipse (blue dashed line) circumscribes the landscape of SARS-CoV-2 NAbs elicited. (B) The distribution of *IIP*_100_ values of NAbs. Average *IIP*_100_ values are used for the 8 NAbs mentioned above.

The landscape contains potent NAbs, with low *IC*_50_ and high *m*, as well as weak NAbs, with the opposite traits. To compare the NAbs, we employed the instantaneous inhibitory potential (*IIP*), a composite metric of *IC*_50_ and *m* (*12-14*). *IIP*_*D*_ represents the log_10_ decline in viral load in a single round infection assay due to the NAb present at concentration *D*. Thus, the higher is the *IIP*_*D*_, the more potent is the NAb at concentration *D*. NAbs displayed a wide distribution of *IIP*_100_ values (Fig. 2B and table S1): We found that 5 NAbs had the highest *IIP*_100_ values, >5, and 9 had the least, <1 (*D* = 100 µg/mL) (Fig. 2B and table S1). This distribution of *IIP*_100_ values demonstrated further the wide spectrum of neutralization efficiencies of NAbs.

The landscape established bounds on the neutralization efficiencies of the NAbs elicited. We reasoned next that the diversity of the NAb responses across individuals would arise from the way NAbs are sampled from the landscape. Although a large number of NAbs can be isolated from individuals, studies of convalescent patient plasma (*5, 6, 16-18*) as well as on NAb epitope profiling (*19*) have argued that the NAb response of an individual can be attributed to a small subset of 5-10 distinct NAbs. Furthermore, while some epitopes on the SARS-CoV-2 spike protein, S, are targeted more than others by NAbs, the collection of NAbs produced differs substantially across individuals (*20*). We therefore assumed that the response elicited by an infected individual would be a *small, random subset* of the landscape. We analysed DRCs of NAbs isolated from individual patients and found that they indeed constituted such random subsets in the landscape (fig. S5). Accordingly, we sampled random combinations of 10 NAbs each, each combination representing the response of an individual. We let NAb concentrations vary across individuals, to mimic the observed variation of the NAb titres (*16-18*). We quantified the neutralization efficiency of the NAb response by simulating standard plasma dilution assays (materials and methods, Fig. 3A). We let the NAbs exhibit Bliss independence or Loewe additivity, the former representing NAbs targeting distinct, non-occluding epitopes and the latter the same or occluding epitopes (*21*). Our simulations recapitulated the dilution curves associated with patient plasma (Fig. 3, B and C). The values of *NT*_50_, the dilution at which the neutralization efficiency of the plasma decreases by 50%, were in agreement with experimental observations (*17*) (Fig. 3D). The data was described better by Bliss independence at low NAb titres and Loewe additivity at high titres. This is expected because at low titres, the NAbs are unlikely to interact with each other and would thus follow Bliss independence, whereas at high titres, they may compete for binding sites on S or occlude each other and thus exhibit Loewe additivity (*21*). At any NAb titre, there existed substantial variation in *NT*_50_, attributed to the random combinations of NAbs sampled. The variation, however, was outweighed by the overall rise of *NT*_50_ with the NAb titre, consistent with patient data (Fig. 3D). For instance, the *NT*_50_ was 17±13 at the IgG titre of 0.1 µg/ml and 1300±1000 at 10 µg/ml. Sampling from the NAb landscape thus successfully recapitulated patient responses. We were able to describe the diversity of the NAb responses elicited across patients. Armed with this description, we examined next the protection accorded by vaccines in clinical trials.

**Figure 3.**
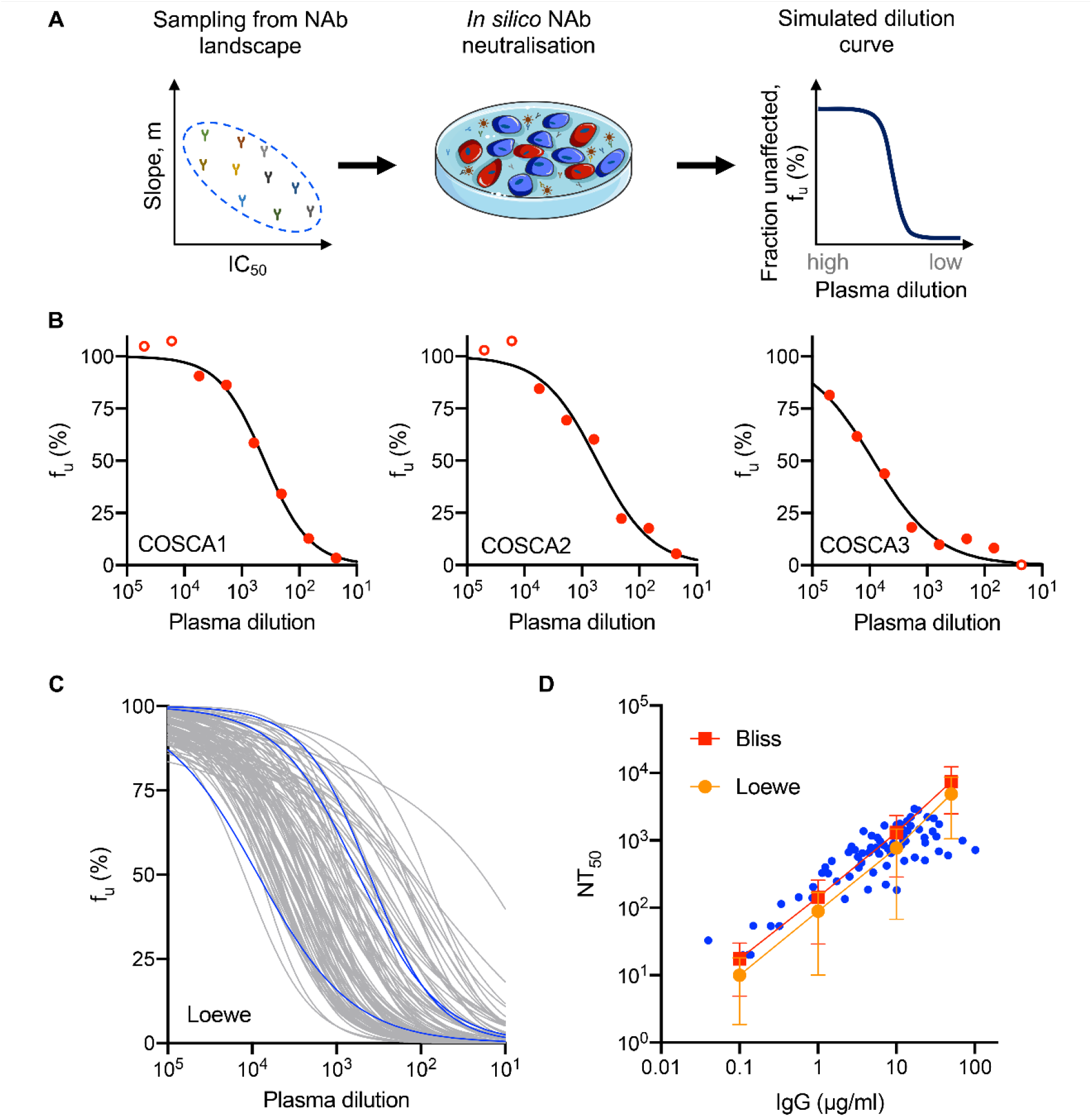
NAb landscape and patient responses. (A) Schematic of the procedure to predict plasma dilution curves. We represent an individual’s plasma by a sample of NAbs from the landscape. We predict the fraction of infection events unaffected by NAbs, *f*_*u*_, at a given plasma dilution in *in vitro* pseudovirus neutralization assays and repeat this at different dilutions to obtain the dilution curve. (B) Representative plasma dilution curves obtained as fits (lines) of the equation 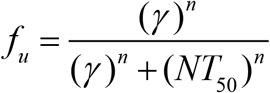 to reported data (circles) from three patients (*15*), where *n* is the Hill coefficient, *γ* is the plasma dilution and *NT*_50_ is the half-maximal inhibitory plasma neutralizing titre. Experimental data points with 1% < *f*_*u*_ < 99% (filled circles) were considered for parameter estimation. (C) Predictions (lines) of plasma dilution curves. We assumed ten NAbs per patient. Blue lines are fits shown in B. *D*_0_ = 30 µg/ml. (D) Half-maximal inhibitory plasma neutralizing titre, *NT*_50_, as a function of total NAb concentration. Blues circles are reported estimates from convalescent patients. Red squares and orange circles are the mean of *NT*_50_ values predicted from 100 virtual patients at each NAb concentration using Bliss independence and Loewe additivity, respectively. The error bars are standard deviations.

Following vaccination, NAb titres rise and are expected to remain stable (or decay slowly) over weeks to months (*22*), protecting individuals who might get exposed to the virus during this period. Individuals were assumed to be protected if they did not report symptomatic infection; loss of protection involved symptoms and a positive result on a nucleic acid amplification test (*1, 2*). Protection with NAbs is expected not to be sterilizing, as suggested by animal studies (*9*); NAbs help suppress the peak in viremia, thereby reducing symptoms, and facilitate more rapid clearance of the infection. If the peak is sufficiently suppressed, no symptoms may result, as is the case with the ∼40% of natural infections that remain asymptomatic (*7*). Here, we assumed that an individual would be detected as symptomatically infected if the viral load rose above a threshold during the infection.

To estimate the peak viral load, we developed a mathematical model of the early time course of the infection, where the viral load typically rises, attains a peak, and declines (*23*), and applied it to describe the effect of vaccination (Fig. 4A, table S2, materials and methods). The structure of the model mimics recent models that have captured patient data of viral load changes following primary infection (*24, 25*) (see Fig. 4B, ε=0). In addition, we assumed that NAbs generated following vaccination would exist at the start of infection and neutralize free viruses, effectively reducing viral infectivity. The greater the reduction in infectivity, the lower the peak viral load (Fig. 4B, ε>0). Significant *de novo* NAb production post-infection typically occurs after the peak in viremia (*7*). We therefore considered pre-existing NAbs as responsible for protection and assumed their titres not to vary substantially during the course of the infection, given the typically short course of the infection and the much longer durability of the NAb response to vaccination (*22*). (Our model is not applicable to natural infection before vaccination; no models are currently capable of correctly describing NAb responses following primary infection.) We let the pre-existing NAbs be drawn as random subsets from the landscape, as we did above. The NAbs neutralized free viruses with an efficiency that we estimated using Loewe additivity between the individual NAbs (Fig. 4). NAb titres in the lung airways are expected to be similar to those in the blood given the close coupling between the lungs and the circulatory system (*7*). We simulated a virtual patient population of 3500 individuals, on the order of the number of individuals infected in the placebo arms of clinical trials. The individuals all had distinct viral dynamics parameters drawn from known ranges (table S2), to mimic interpatient variability in addition to the variability arising from NAb sampling from the landscape. Our model predicted wide variability in the peak viral load (Fig. 4C). At low pre-existing NAb concentrations (0.01 µg/mL), indicative of the scenario without vaccination, the predicted peak viral load ranged from ∼10^3^ to 10^9^ copies/ml, consistent with the range in symptomatic individuals (*26*). The peaks declined as NAb titres increased. The limit of detection is ∼10^2^ copies/ml (*27*), which we set as the threshold for symptomatic infection that would be detected in trials. The fraction of individuals with peaks below detection would indicate the level of protection due to the vaccine.

**Figure 4.**
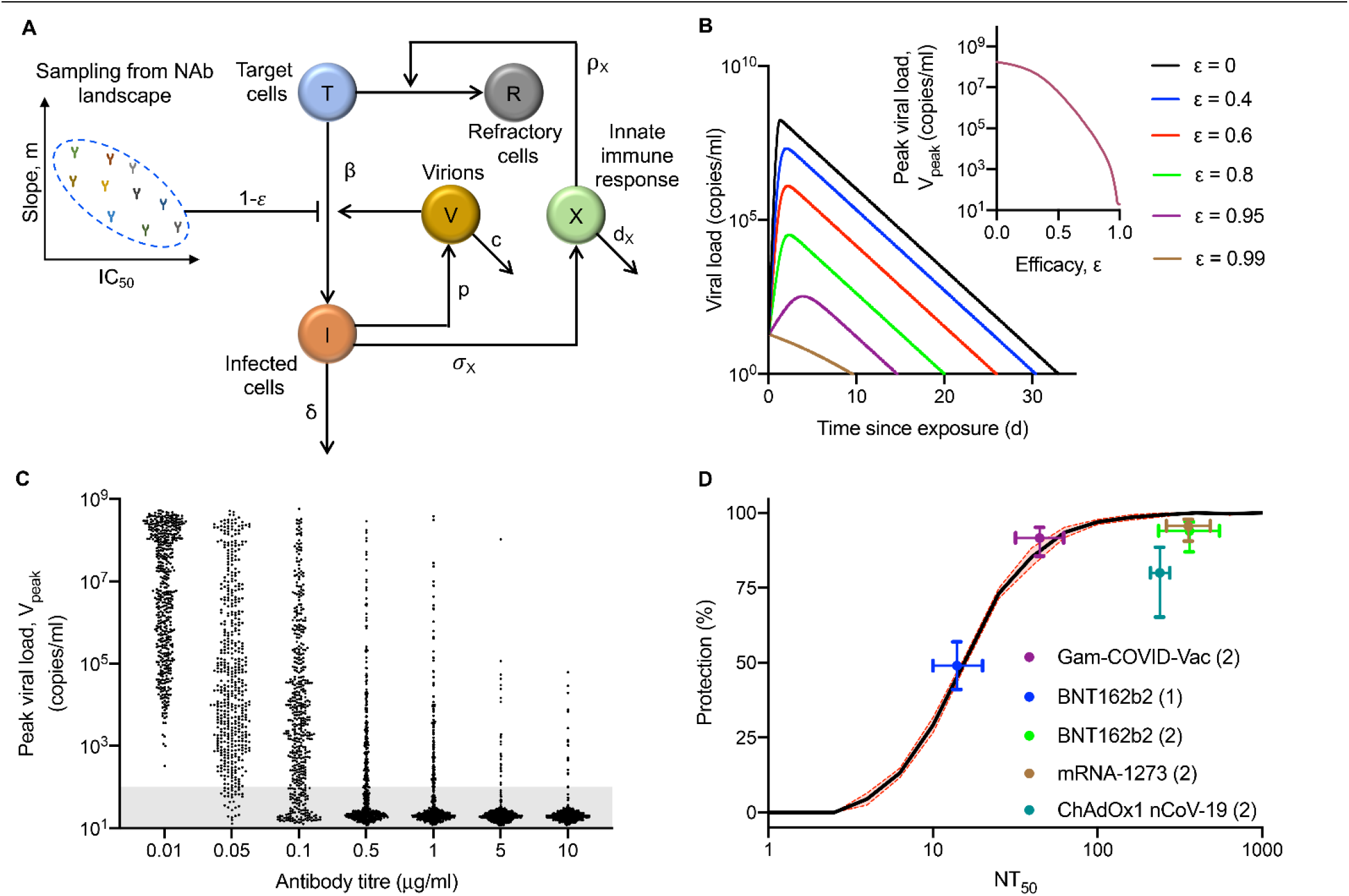
SARS-CoV-2 dynamics and protection post-vaccination. (A) Schematic of the model of within-host SARS-CoV-2 dynamics post-vaccination depicting the interactions between target cells, *T*, infected cells, *I*, refractory cells, *R*, virions, *V*, innate immune response, *X*, and pre-existing NAbs, sampled from the landscape. (B) Predictions of viral load in non-vaccinated (black line) and vaccinated (coloured lines) individuals with different fixed efficacies of NAbs indicated. *Inset*: Predicted peak viral load at different efficacies. (C) Predictions of peak viral load at different NAb titres. Each dot represents a patient. (D) Model predictions of the relationship between mean protection and *NT*_50_ (solid line) compared with data from vaccination trial (symbols). The number of doses of the vaccine administered is mentioned in brackets. The error bars (dashed lines) in the protection curve are the standard deviation from 5 realizations of *in silico* patient populations. The data from the trials used is summarized in table S3. The model equations and simulation procedure are described in materials and methods.

To quantify the mean level of protection and test it against data from clinical trials, we used viral dynamics parameters representative of symptomatic infections (*24, 25*) (table S2) and simulated the dynamics in 5 cohorts of 2000 infected individuals each. Vaccination studies report the *NT*_50_ values of the NAb responses elicited and the associated mean protection level, or efficacy (table S3). We binned the different individuals into narrow *NT*_50_ bands and calculated the mean protection in each band. We found that the mean protection was low for *NT*_50_∼1. It increased in a sigmoidal manner to 50% at *NT*_50_∼20 and asymptotically reached 100% at *NT*_50_∼200. Remarkably, the data for nearly all approved vaccines fell on this ‘protection curve’, explaining the protection they confer (Fig. 4D). Thus, for instance, a single dose of the vaccine BNT 162b2 elicited NAbs with *NT*_50_ of 14 and accorded 49% protection. Following two doses, the corresponding values were 361 and 94%, respectively. These values as well as those for other vaccines were captured accurately by our model predictions. The only exception was ChAdOx1 nCoV-19, which had a lower protection than predicted, the reasons for which remain to be elucidated.

Our study provides the first conceptual, mechanistic and quantitative understanding of the protection conferred by COVID-19 vaccines. Our findings would inform strategies for optimal vaccine deployment. With limited vaccine availability, it would be useful to estimate the protection realizable by a single dose of a prime-boost vaccine, especially in younger, less vulnerable adults (*10*). Our formalism would enable this estimation: measurements of corresponding *NT*_50_ values would allow reading off the expected protection levels from our protection curve. Similarly, using measurements of the waning of NAb titres post-vaccination, how the population-level protection due to pre-existing NAbs would fade could be predicted. Protection would then rely on memory B cell responses, which are yet to be fully understood (*28*), or indicate the need for revaccination. Our study did not consider viral mutations because with 5-10 NAbs active, viral escape from NAb responses is expected to be unlikely (*19, 29*). With the new circulating strains (*30*), however, the NAb landscape may have to be reconstructed. Future studies may report DRCs of NAbs against the new strains, facilitating such reconstruction.

## Supporting information

SI combined

## Data Availability

All data is available within the submission.

## Author contributions

PP: Conceptualization, Investigation, Formal analysis, Writing-original draft, Writing-review & editing; RD: Formal analysis, Writing-review & editing. NMD: Conceptualization, Writing-original draft, Writing-review & editing.

## Data and materials availability

All relevant data are available within the manuscript and supplementary materials.

