## Supplementary material for "Modelling the population-level protection conferred by COVID-19 vaccination": SI combined

**This PDF file includes:**

Materials and Methods
Figs. S1 to S5
Tables S1 to S3

#### Materials and Methods

##### *Data*

We considered data from studies that reported *in vitro* dose-response curves of NAbs using SARS-CoV-2 pseudotyped virions (*5, 6, 31-47*). The assays estimated the fraction of infection events unaffected by the NAbs as a function of the NAb concentration (Fig. 1, and figs. S2 and S3). Data from such assays have been successfully used to estimate $m$ and $IC_{50}$ of antibodies against HIV-1 (*12*) and HCV (*13*). We extracted the data using Engauge Digitizer 12.1 and ensured consistency with reported details, such as dilution levels used.

##### *Analysis of DRCs*

We used both the standard sigmoidal dose-response curve equation (Eq. [1]) and the median-effect equation (Eq. [2]) to analyse the data.

$$f_u = 1 - f_a = \frac{(IC_{50})^m}{(D)^m + (IC_{50})^m} \tag{1}$$

$$\log_{10}\left(\frac{f_a}{f_u}\right) = m\log_{10}\left(\frac{D}{IC_{50}}\right) \tag{2}$$

Here, $f_u$ and $f_a$ are the fraction of infection events unaffected and affected by the NAbs in a single round of infection, $D$ is the NAb concentration, $IC_{50}$ is the half-maximal inhibitory concentration and $m$ is the slope. Data was fitted using the tool NLINFIT in MATLAB R2017b. Data points with $1\% < f_u < 99\%$ were considered for parameter estimation. We fit the data using Eq. [1] and Eq. [2] separately and obtained estimates of $IC_{50}$ and $m$ for each NAb as well as associated 95% confidence intervals. We then computed

$IIP_{100} = \log_{10}\left(1+\left(\frac{100}{IC_{50}}\right)^{m}\right)$ using the estimates obtained using Eq. [1] and Eq. [2]. In most cases, the $IIP_{100}$ values were close to each other. We did not include NAbs for which $IIP_{100}$ values estimated using the two methods deviated by 20% or more in our analysis (table S1), for the deviation indicated that such NAbs either did not conform to the trends expected by Eqs. [1] and [2] or had large uncertainties in the data precluding robust parameter estimation. The details of the NAbs and parameter estimates are presented in table S1.

***In silico simulation of plasma dilution assays***

We simulated plasma dilution experiments as follows. We assumed that the plasma contained $N$ NAbs with equimolar concentrations sampled from the landscape (Fig. 2A). The reciprocal plasma dilution curve was predicted assuming Loewe additivity (Eq. 3) or Bliss independence (Eq. [4]) between the different NAbs (*48-50*) using

$$\sum_{i=1}^{N} \frac{D_i / g}{IC_{50_i}\left(\frac{1}{e_L} - 1\right)^{-1/m_i}} = 1 \qquad (3)$$

$$e_B = 1 - \prod_{i=1}^{N} \frac{\left(IC_{50_i}\right)^{m_i}}{\left(IC_{50_i}\right)^{m_i} + \left(D_i / g\right)^{m_i}} \qquad (4)$$

Here, $g$ is the plasma dilution factor. $e_L$ and $e_B$ are the fractions of infection events affected by the plasma in a single round of infection estimated using Loewe additivity and Bliss independence, respectively. $D_i$ is the concentration of the $i^{th}$ NAb in the plasma before dilution, $IC_{50_i}$ is its half-maximal inhibitory concentration and $m_i$ its slope, with

$i \in \{1,2,..,N\}$. We let $N$ = 10 in our simulations, based on the number of NAbs with significant neutralization efficacy seen in patients (*5*). We estimated the value of $\gamma$ at which $\varepsilon = 0.5$ as the corresponding $NT_{50}$. We chose $D_i$ as $D_0 /N$, and varied $D_0$ between 0.1 and100 µg/ml.

We repeated these simulations 100 times at different NAb titres, with each simulation representative of an individual patient. We compared the resulting predictions at $D_0$ = 30 µg/ml with observations from 3 patients (Fig. 3C) (*15*), which also we digitized (Fig. 3B). The equation $f_u = \frac{(\gamma)^n}{(\gamma)^n + (NT_{50})^n}$ was fit to the observations from 3 patients. Here, $n$ is the Hill coefficient, $\gamma$ is the plasma dilution and $NT_{50}$ is the half-maximal inhibitory plasma neutralizing titre. We also compared predictions of the dependence of $NT_{50}$ on NAb titres with experimental observations (Fig. 3D) (*17*).

##### *Model of SARS-CoV-2 dynamics post-vaccination*

To predict the protection conferred by vaccines, we developed a mathematical model of within-host SARS-CoV-2 infection post-vaccination. We adapted previous models (*24, 51-54*) by focussing on early dynamics, required to accurately predict the reduction in the peak viral load due to pre-existing NAbs. The following equations described the resulting infection dynamics in vaccinated individuals exposed to the virus:

$$\frac{dT}{dt} = -\beta(1-\varepsilon)VT - \rho_X XT \quad (5)$$

$$\frac{dR}{dt} = \rho_X XT \quad (6)$$

$$\frac{dI}{dt} = b(1 - e)VT - dI \tag{7}$$

$$\frac{dV}{dt} = pI - cV \tag{8}$$

$$\frac{dX}{dt} = \frac{s_X I(1 - X)}{f_X + I} - d_X X \tag{9}$$

Here, uninfected target cells, *T*, are infected by SARS-CoV-2 virions, *V*, with the second order rate constant $b$, producing infected cells, *I*. Cells *I* produce virions at the rate $p$ per cell and are lost with a rate constant $d$. The virions are cleared with a rate constant $c$. The activation of the innate immune response, quantified phenomenologically using *X*, is assumed to be a saturable function of *I*, with the maximal rate $s_X$ and the half-maximal activation parameter $f_X$. If *I* is not limiting, *X* would rise at the rate $s_X$ at low *X* and cease to rise as *X* approaches 1. *X* converts uninfected cells to an infection-refractory state, *R*, at a per capita rate $r_X$ and decays with a rate constant $d_X$. The pre-existing NAbs are drawn as random subsets from the landscape (Fig. 2A) to block new infections with an efficacy $e$, which is a function of NAb titre and computed using Eq. [1] or Eq. [2].

To examine the variation in peak viral loads with NAb titres (Fig. 4C), we predicted viral dynamics of 3500 infected individuals, each individual with different pre-existing NAbs sampled from the landscape as well as different values of the viral dynamics parameters (table S2). To obtain the protection curve (Fig. 4D), we simulated viral dynamics in 2000 virtual individuals, as we described above, and obtained peak viral loads. Further, for each individual, we also simulated plasma dilution assays and estimated $NT_{50}$. We then binned individuals into narrow ranges of $NT_{50}$ values. In each bin, we estimated the fraction of individuals with peak viral load below 100 copies/ml. This fraction yielded the protection curve. The error bars (standard deviation) of the protection curve were obtained from 5 independent realizations.

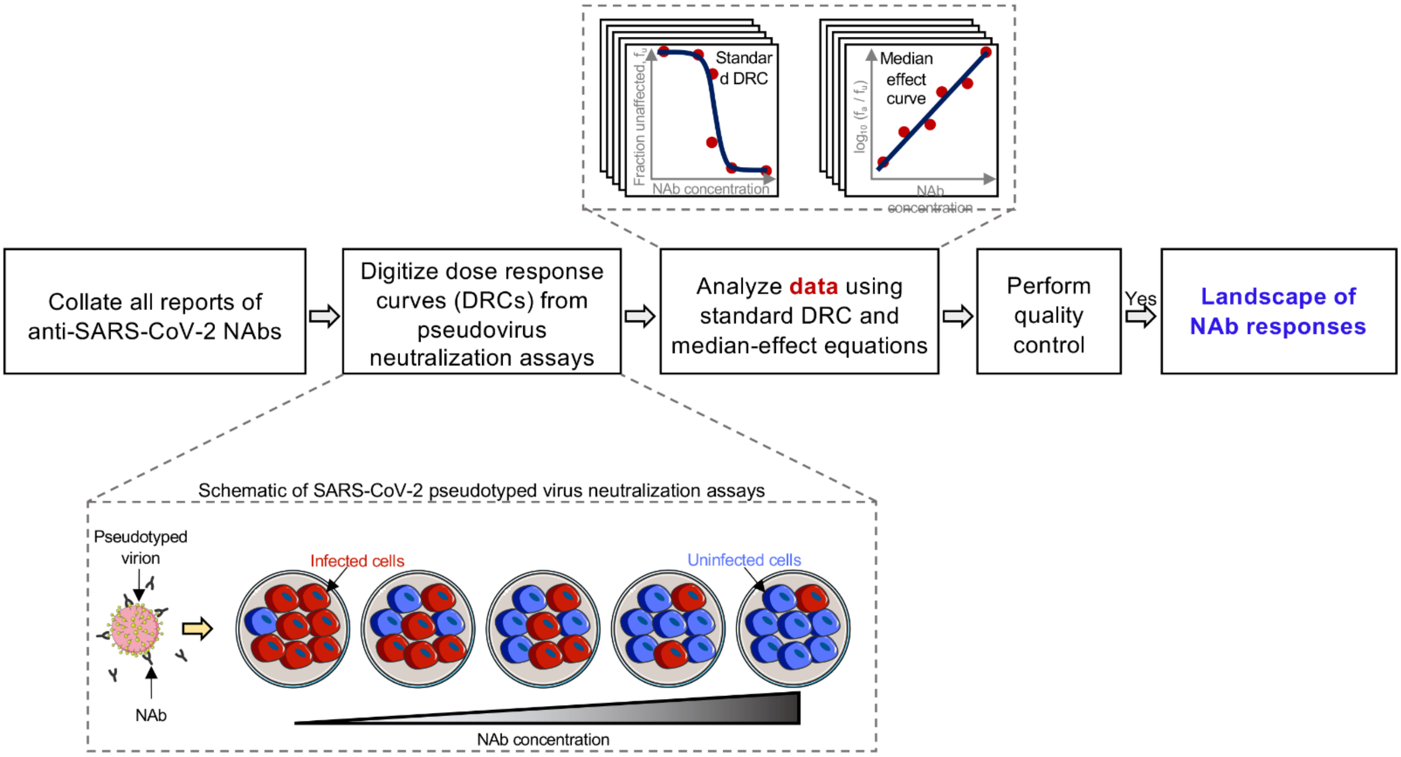


**Fig. S1. Schematic of the workflow to chart out the quantitative landscape of SARS-CoV-2 NAbs.** We collated data from a large number of studies that reported DRCs of NAbs using SARS-CoV-2 pseudotyped virions. The assays estimate the fraction of infection events affected/unaffected by the NAbs as a function of the NAb concentration. We extracted and analysed the data using both the standard DRC equation (Eq. [1]) and the median-effect equation (Eq. [2]) to estimate $IC_{50}$ and $m$. We then computed $IIP_{100} = \log_{10}\left(1+\left(\frac{100}{IC_{50}}\right)^m\right)$ using the estimates obtained by both equations. NAbs with consistent estimates were considered for rank-ordering.

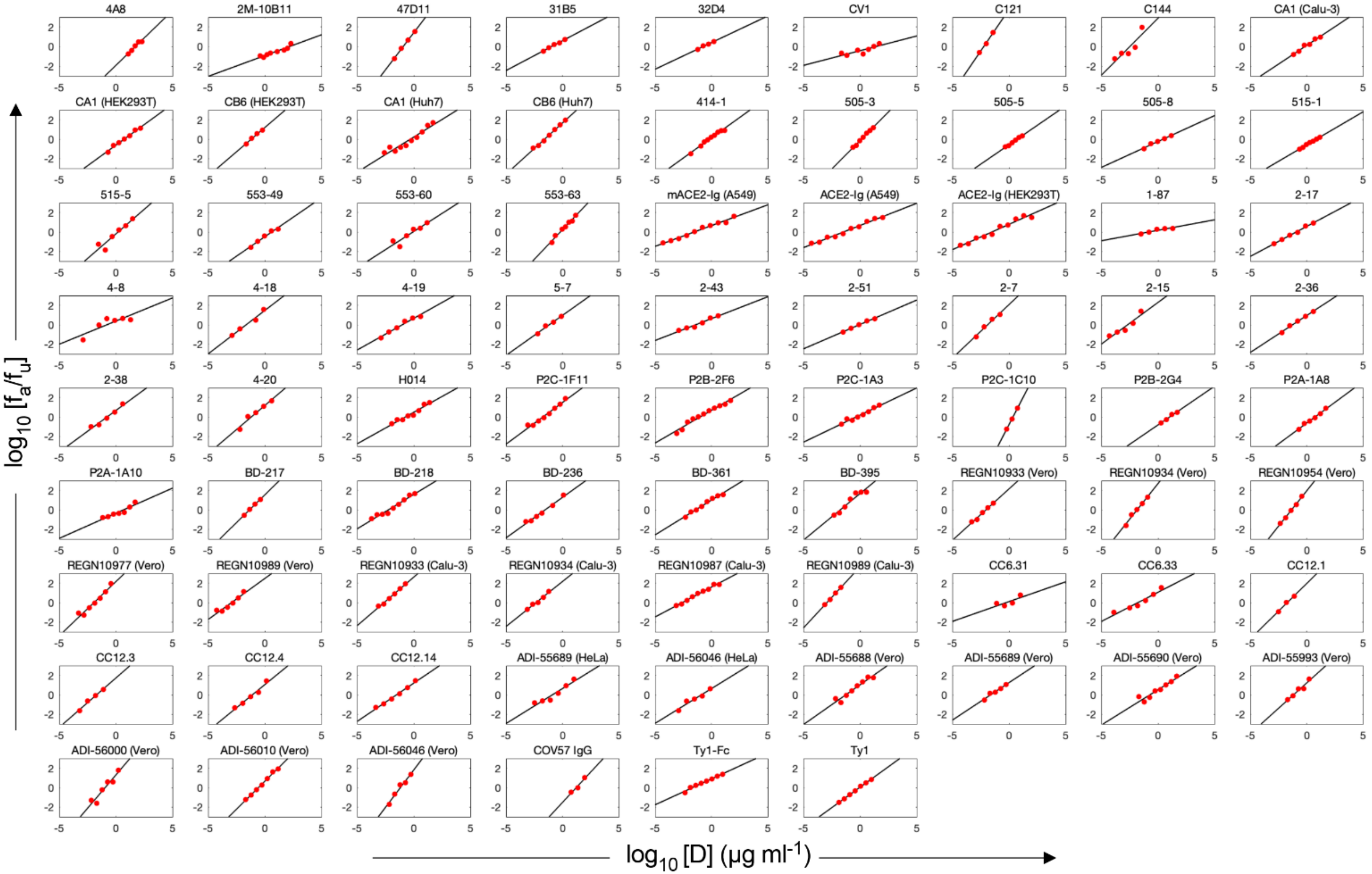


**Fig. S2. Estimates of $IC_{50}$ and $m$ of SARS-CoV-2 NAbs using the median-effect equation.** Fits (lines) of the median-effect equation to published experimental data (circles). Experimental data points with 1% < $f_u$ < 99% (filled circles) were considered for parameter estimation.

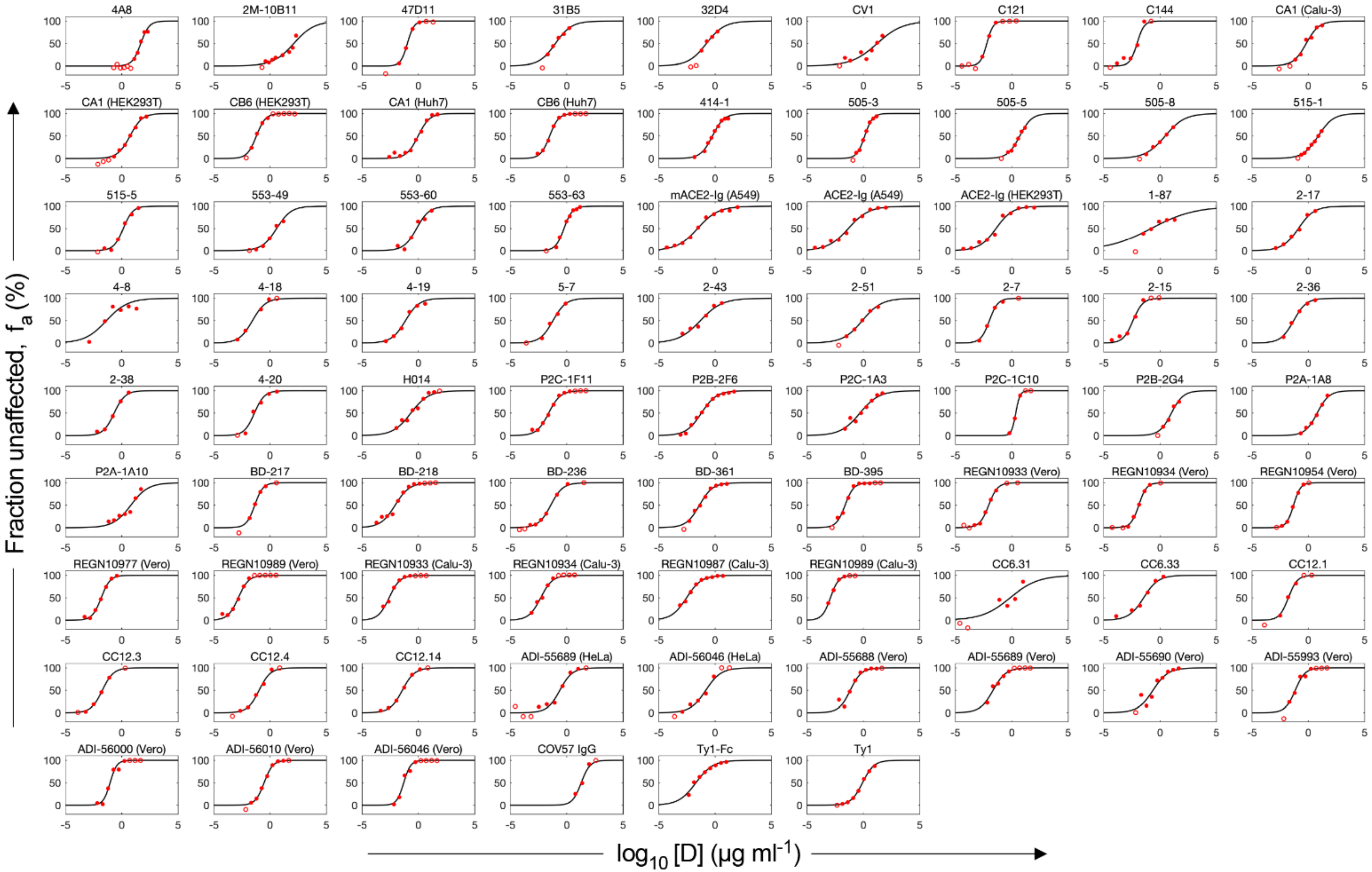


**Fig. S3. Estimates of $IC_{50}$ and $m$ of SARS-CoV-2 NAbs using the standard dose-response curve equation.** Fits (lines) of the standard dose-response curve equation to published experimental data (circles). Experimental data points with $1\% < f_u < 99\%$ (filled circles) were considered for parameter estimation.

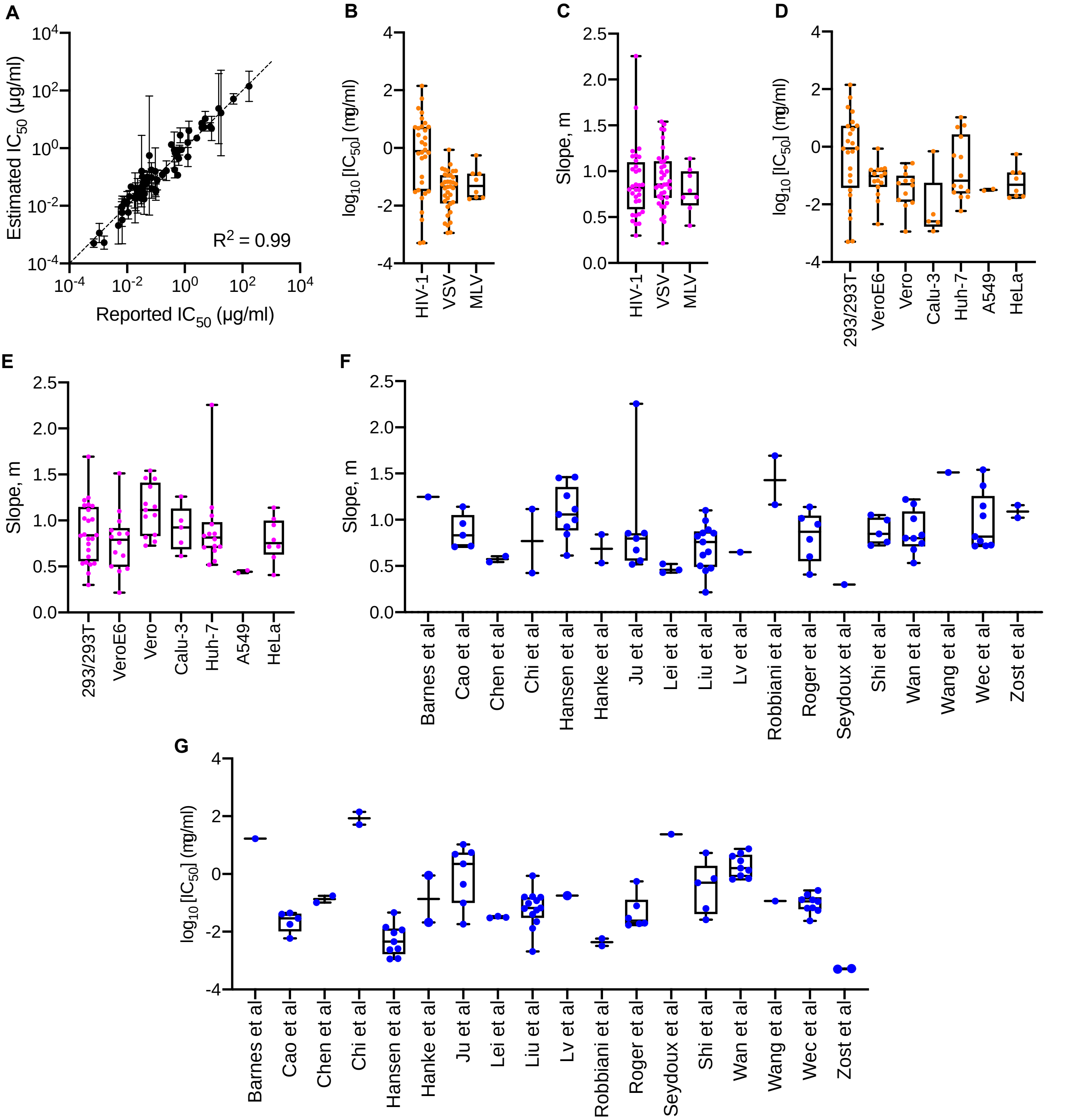


**Fig. S4. Robustness of the estimates of $IC_{50}$ and $m$ of SARS-CoV-2 NAbs.** (A) Comparison of the estimated and reported values of $IC_{50}$ (see table S1). Error bars are 95% confidence intervals. (B, C) Variability in $m$ (B) and $IC_{50}$ (C) within different studies (see table S1 for details of the studies).

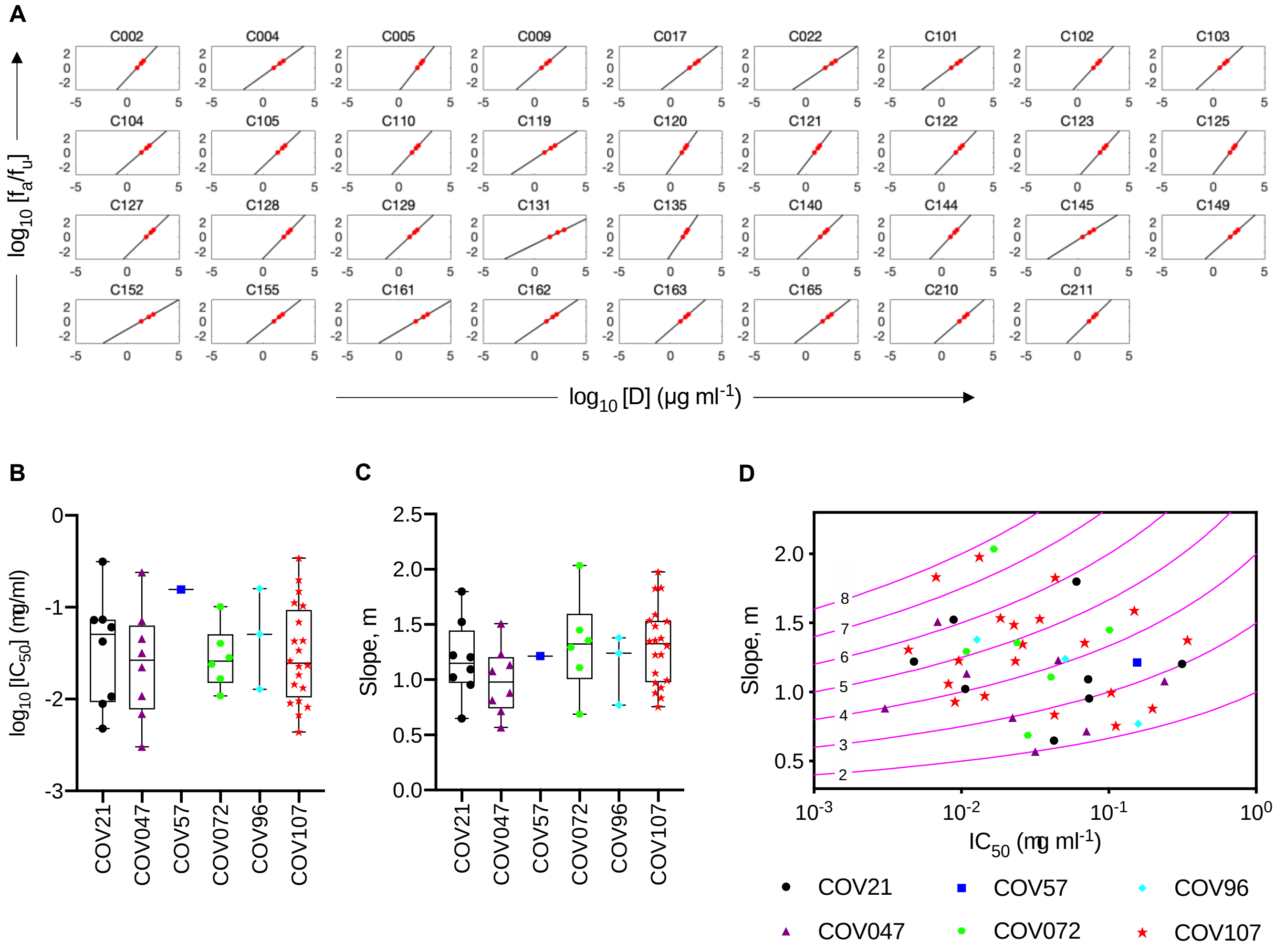

**Fig. S5. Estimates of $IC_{50}$ and $m$ of SARS-CoV-2 NAbs from patients. (A)** Fits (lines) of the median-effect equation to published experimental data (circles) reporting $IC_{50}$, $IC_{80}$ and $IC_{90}$ for NAbs drawn from eight patients (*6*). For some NAbs, only $IC_{50}$ and $IC_{80}$ were available. The slopes for these latter NAbs were estimated using the equation $m = \frac{\log_{10}(4)}{\log_{10}(IC_{80}) - \log_{10}(IC_{50})}$ derived from the median-effect equation (Eq. [2]). The corresponding best-fit estimates of (B) $IC_{50}$ and (C) $m$ are shown along with (D) the location of the NAbs on the landscape.

**Table S1. Best-fit estimates of dose-response curve parameters of NAbs.**

| Data | | | | | Fits and predictions | | | | | |
|---|---|---|---|---|---|---|---|---|---|---|
| | | | | | Median-effect equation | | | Standard dose response curve | | |
| Antibody | Cell line | Viral backbone | Reported $IC_{50}$ | Ref | m (95% CI) | $IC_{50}$ (95% CI) | $IIP_{100}$ | m (95% CI) | $IC_{50}$ (95% CI) | $IIP_{100}$ |
| 4A8 | 293T/ACE2 | HIV-1 | 49 μg/ml | (*31*) | 1.12 (0.64, 1.59) | 51.29 μg/ml (33.75, 77.96) | 0.49 | 1.17 (0.61, 1.73) | 48.06 μg/ml (33.09, 69.81) | 0.53 |
| 2M-10B11 | 293T/ACE2 | HIV-1 | 170 μg/ml | (*31*) | 0.42 (0.28, 0.57) | 140.51 μg/ml (41.89, 471.27) | 0.27 | 0.48 (0.26, 0.7) | 126.72 μg/ml (52.48, 305.96) | 0.28 |
| 47D11 | VeroE6 | VSV | 0.57 μg/ml | (*32*) | 1.51 μg/ml (1.29, 1.74) | 0.12 μg/ml (0.09, 0.15) | 4.44 | 1.48 (1.15, 1.81) | 0.11 μg/ml (0.09, 0.13) | 4.39 |
| 31B5 | 293T/ACE2 | - | 0.034 μg/ml | (*46*) | 0.61 (0.48, 0.73) | 0.1 μg/ml (0.07, 0.14) | 1.81 | 0.6 (0.45, 0.76) | 0.1 μg/ml (0.07, 0.14) | 1.82 |
| 32D4 | 293T/ACE2 | - | 0.07 μg/ml | (*46*) | 0.54 (0.31, 0.77) | 0.18 μg/ml (0.1, 0.32) | 1.51 | 0.55 (0.27, 0.82) | 0.17 μg/ml (0.09, 0.31) | 1.52 |
| CV1 | 293T/ACE2 | HIV-1 | 15 μg/ml | (*33*) | 0.3 (0.08, 0.52) | 23.67 μg/ml (1.44, 390.2) | 0.4 | 0.38 (0.11, 0.66) | 16.96 μg/ml (3.88, 74.12) | 0.47 |
| CV30[$] | 293T/ACE2 | HIV-1 | 0.03 μg/ml | (*33*) | 0.91 (0.51, 1.31) | 0.03 μg/ml (0.01, 0.1) | 3.17 | 1.19 (0.7, 1.68) | 0.04 μg/ml (0.03, 0.06) | 4.05 |
| C121[#] | 293T/ACE2 | HIV-1 | 6.7 ng/ml | (*6*) | 1.69 (0.19, 3.2) | 5.81 ng/ml (1.92, 17.56) | 7.17 | 1.54 (0.17, 2.91) | 6.09 ng/ml (3.18, 11.65) | 6.51 |
| C135[$#] | 293T/ACE2 | HIV-1 | 16.5 ng/ml | (*6*) | 0.87 (0.39, 1.35) | 21.65 ng/ml (4.93-95.12) | 3.2 | 1.91 (1.08, 2.75) | 16.35 ng/ml (12.13, 22.03) | 7.24 |

| | | | | | | | | | | |
|---|---|---|---|---|---|---|---|---|---|---|
| C144*# | 293T/ACE2 | HIV-1 | 6.8 ng/ml | (*6*) | 1.16 (0.04, 2.28) | 3.24 ng/ml (0.48, 21.74) | 5.22 | 1.41 (0, 3.09) | 9 ng/ml (3.64, 22.23) | 5.69 |
| CA1 | Calu-3 | HIV-1 | 0.53 µg/ml | (*34*) | 0.76 (0.58, 0.94) | 0.69 µg/ml (0.44, 1.09) | 1.65 | 0.77 (0.47, 1.07) | 0.64 µg/ml (0.39, 1.07) | 1.69 |
| CA1 | 293T/ACE2 | HIV-1 | 4.66 µg/ml | (*34*) | 0.85 (0.72, 0.97) | 5.4 µg/ml (3.93, 7.41) | 1.11 | 0.8 (0.68, 0.92) | 5.22 µg/ml (4.26, 6.4) | 1.07 |
| CA1 | Huh-7 | HIV-1 | 1.28 µg/ml | (*34*) | 0.72 (0.55, 0.9) | 0.5 µg/ml (0.23, 1.06) | 1.68 | 0.85 (0.64, 1.07) | 0.95 µg/ml (0.68, 1.31) | 1.74 |
| CB6$* | Calu-3 | HIV-1 | 0.02 µg/ml | (*34*) | 1.33 (0.37, 2.28) | 0.04 µg/ml (0.01, 0.13) | 4.5 | 1.68 (0, 3.59) | 0.03 µg/ml (0.01, 0.06) | 6 |
| CB6 | 293T/ACE2 | HIV-1 | 0.04 µg/ml | (*34*) | 1 (0.66, 1.33) | 0.06 µg/ml (0.04, 0.1) | 3.19 | 1.07 (0.68, 1.45) | 0.06 µg/ml (0.04, 0.08) | 3.44 |
| CB6 | Huh-7 | HIV-1 | 0.04 µg/ml | (*34*) | 1.05 (0.95, 1.15) | 0.03 µg/ml (0.02, 0.03) | 3.77 | 1.09 (0.93, 1.26) | 0.03 µg/ml (0.03, 0.04) | 3.85 |
| 414-1 | 293T/ACE2 | HIV-1 | 3.09 nM | (*47*) | 0.83 (0.73, 0.94) | 4.54 nM (3.49, 5.91) | 1.81 | 0.83 (0.72, 0.95) | 3.89 nM (3.31, 4.58) | 1.87 |
| 505-3 | 293T/ACE2 | HIV-1 | 2.31 nM | (*47*) | 1.17 (1.06, 1.29) | 8.82 nM (7.67, 10.14) | 2.2 | 1.25 (1.1, 1.39) | 8.71 nM (7.88, 9.64) | 2.35 |
| 505-5 | 293T/ACE2 | HIV-1 | 26.37 nM | (*47*) | 0.8 (0.68, 0.92) | 34.67 nM (28.06, 42.84) | 1.07 | 0.82 (0.69, 0.95) | 34.1 nM (29.06, 40) | 1.09 |
| 505-8 | 293T/ACE2 | HIV-1 | 4.67 nM | (*47*) | 0.53 (0.39, 0.67) | 18.73 nM (10.35, 33.88) | 0.89 | 0.5 (0.38, 0.62) | 18.63 nM (12.68, 27.39) | 0.84 |
| 515-1 | 293T/ACE2 | HIV-1 | 26.11 nM | (*47*) | 0.68 (0.61, 0.74) | 48.56 nM (40.4, 58.35) | 0.84 | 0.65 (0.59, 0.72) | 49.49 nM (43.37, 56.47) | 0.81 |

| | | | | | | | | | | |
|---|---|---|---|---|---|---|---|---|---|---|
| 515-5 | 293T/ACE2 | HIV-1 | 5.39/8.47 nM | (*47*) | 1.01 (0.53, 1.5) | 10.54 nM (3.3, 33.67) | 1.83 | 1.04 (0.71, 1.37) | 9.01 nM (6.41, 12.65) | 1.95 |
| 553-49 | 293T/ACE2 | HIV-1 | 9.26 nM | (*47*) | 0.8 (0.55, 1.05) | 27.13 nM (12.68, 58.04) | 1.15 | 0.68 nM (0.36, 1.01) | 25.35 (13.23, 48.59) | 1.01 |
| 553-60 | 293T/ACE2 | HIV-1 | 2.87 nM | (*47*) | 0.75 (0.31, 1.18) | 5.78 nM (1.37, 24.38) | 1.55 | 0.78 (0.34, 1.21) | 4.64 nM (2.13, 10.11) | 1.69 |
| 553-63 | 293T/ACE2 | HIV-1 | 3.57 nM | (*47*) | 1.22 (1.02, 1.42) | 4.32 nM (3.18, 5.85) | 2.67 | 1.17 (0.93, 1.41) | 3.8 nM (3.06, 4.73) | 2.63 |
| mACE2-Ig | A549/ACE2 | HIV-1 | 0.03 μg/ml | (*36*) | 0.43 (0.38, 0.47 | 0.03 μg/ml (0.02, 0.05) | 1.52 | 0.47 (0.41, 0.54) | 0.03 μg/ml (0.02, 0.04) | 1.69 |
| mACE2-Ig[$] | 293T/ACE2 | HIV-1 | 0.08 μg/ml | (*36*) | 0.55 (0.44, 0.67 | 0.02 μg/ml (0.01, 0.05) | 2.07 | 0.7 (0.53, 0.86) | 0.02 μg/ml (0.02, 0.04) | 2.52 |
| ACE2-Ig | A549/ACE2 | HIV-1 | 0.1 μg/ml | (*36*) | 0.46 (0.41, 0.51 | 0.03 μg/ml (0.02, 0.06) | 1.59 | 0.48 (0.38, 0.57) | 0.05 μg/ml (0.03, 0.07) | 1.6 |
| ACE2-Ig | 293T/ACE2 | HIV-1 | 0.1 μg/ml | (*36*) | 0.52 (0.44, 0.6) | 0.03 μg/ml (0.02, 0.06) | 1.83 | 0.59 (0.42, 0.77) | 0.04 μg/ml (0.02, 0.07) | 2.02 |
| 2-26[$] | VeroE6 | VSV | - | (*5*) | 0.36 (0.11, 0.62) | 4.42 μg/ml (0.33, 60.08) | 0.61 | 0.53 (0.16, 0.89) | 3.58 μg/ml (1.03, 12.4) | 0.83 |
| 1-68[$] | VeroE6 | VSV | 0.767 μg/ml | (*5*) | 0.65 (0.001, 1.3) | 1.68 μg/ml (0.03, 109.8) | 1.18 | 0.7 (0.42, 0.98) | 0.95 μg/ml (0.54, 1.66) | 1.43 |
| 1-87 | VeroE6 | VSV | 0.095 μg/ml | (*5*) | 0.22 (0.07, 0.36) | 0.16 μg/ml (0.02, 1.03) | 0.7 | 0.23 (0.08, 0.38) | 0.16 (0.03, 0.82) | 0.72 |
| 2-17 | VeroE6 | VSV | 0.168 μg/ml | (*5*) | 0.62 (0.54, 0.69) | 0.12 μg/ml (0.08, 0.17) | 1.81 | 0.63 (0.47, 0.78) | 0.13 μg/ml (0.08, 0.2) | 1.81 |

| | | | | | | | | | | |
|---|---|---|---|---|---|---|---|---|---|---|
| 4-8* | VeroE6 | VSV | 0.032 μg/ml | (*5*) | 0.48 (0.05, 0.9) | 0.16 μg/ml (0.01, 2.76) | 1.35 | 0.46 (0, 0.92) | 0.04 μg/ml (0.01, 0.42) | 1.55 |
| 4-18 | VeroE6 | VSV | 0.023 μg/ml | (*5*) | 0.89 (0.41, 1.37) | 0.02 μg/ml (0.01, 0.09) | 3.26 | 0.74 (0.35, 1.13) | 0.03 μg/ml (0.01, 0.08) | 2.64 |
| 4-19 | VeroE6 | VSV | 0.07 μg/ml | (*5*) | 0.65 (0.49, 0.81) | 0.1 μg/ml (0.05, 0.19) | 1.97 | 0.71 (0.48, 0.94) | 0.07 μg/ml (0.04, 0.12) | 2.22 |
| 5-7 | VeroE6 | VSV | 0.05 μg/ml | (*5*) | 0.82 (0.47, 1.17) | 0.07 μg/ml (0.03, 0.14) | 2.61 | 0.76 (0.27, 1.25) | 0.06 μg/ml (0.03, 0.14) | 2.46 |
| 5-24$ | VeroE6 | VSV | 0.013 μg/ml | (*5*) | 0.3 (0.02, 0.58) | 0.005 μg/ml ($1 \times 10^{-4}$, 0.28) | 1.3 | 0.4 (0.13, 0.66) | 0.009 μg/ml (0.002, 0.04) | 1.62 |
| 2-43 | VeroE6 | VSV | 0.071 μg/ml | (*5*) | 0.45 (0.32, 0.58) | 0.04 μg/ml (0.02, 0.09) | 1.54 | 0.45 (0.28, 0.62) | 0.05 μg/ml (0.02, 0.11) | 1.5 |
| 2-51 | VeroE6 | VSV | 0.652 μg/ml | (*5*) | 0.5 (0.42, 0.59) | 0.86 μg/ml (0.58, 1.27) | 1.08 | 0.51 (0.42, 0.6) | 0.8 μg/ml (0.57, 1.12) | 1.11 |
| 1-20$* | VeroE6 | VSV | 0.127 μg/ml | (*5*) | 1.2 (0.18, 2.23) | 0.06 μg/ml (0.01, 0.29) | 3.85 | 0.87 (0, 2.08) | 0.09 μg/ml (0.02, 0.47) | 2.66 |
| 1-57$ | VeroE6 | VSV | 0.009 μg/ml | (*5*) | 0.97 (0.52, 1.42) | 0.01 μg/ml (0.003, 0.03) | 3.86 | 0.72 (0.3, 1.13) | 0.01 μg/ml (0.005, 0.02) | 2.86 |
| 2-4$ | VeroE6 | VSV | 0.394 μg/ml | (*5*) | 1.27 (0.28, 2.26) | 0.58 μg/ml (0.14, 2.46) | 2.84 | 0.92 (0.19, 1.66) | 0.44 μg/ml (0.18, 1.12) | 2.18 |
| 2-7 | VeroE6 | VSV | 0.01 μg/ml | (*5*) | 1.1 (0.57, 1.64) | 0.01 μg/ml (0.005, 0.03) | 4.28 | 1.15 (0.61, 1.68) | 0.01 μg/ml (0.007, 0.02) | 4.57 |
| 2-15 | VeroE6 | VSV | 0.005 μg/ml | (*5*) | 0.86 (0.31, 1.4) | 0.002 μg/ml (0.001, 0.009) | 4.01 | 1.01 (0.3, 1.72) | 0.004 μg/ml (0.002, 0.009) | 4.45 |

| | | | | | | | | | | |
|---|---|---|---|---|---|---|---|---|---|---|
| 2-30$ | VeroE6 | VSV | 0.512 µg/ml | (*5*) | 0.87 (0.37, 1.36) | 0.36 µg/ml (0.06, 2.23) | 2.12 | 1.17 (0.92, 1.41) | 0.46 µg/ml (0.38, 0.56) | 2.73 |
| 2-36 | VeroE6 | VSV | 0.044 µg/ml | (*5*) | 0.76 (0.64, 0.88) | 0.06 µg/ml (0.04, 0.08) | 2.47 | 0.78 (0.58, 0.97) | 0.05 µg/ml (0.04, 0.07) | 2.57 |
| 2-38 | VeroE6 | VSV | 0.232 µg/ml | (*5*) | 0.86 (0.56, 1.15) | 0.17 µg/ml (0.08, 0.36) | 2.38 | 0.9 (0.64, 1.16) | 0.21 µg/ml (0.15, 0.31) | 2.41 |
| 4-20 | VeroE6 | VSV | 0.036 µg/ml | (*5*) | 0.99 (0.6, 1.38) | 0.06 µg/ml (0.02, 0.17) | 3.16 | 0.96 (0.25, 1.67) | 0.04 µg/ml (0.02, 0.09) | 3.27 |
| H014 IgG | Vero | VSV | 3 nM | (*37*) | 0.65 (0.5, 0.8) | 1.18 nM (0.62, 2.23) | 1.79 | 0.55 (0.38, 0.72) | 1.44 nM (0.83, 2.51) | 1.48 |
| COV2-2196 | 293/ACE2 | HIV-1 | 0.7 ng/ml | (*38*) | 1.16 (0.98, 1.33) | 0.5 ng/ml (0.36, 0.7) | 6.13 | 1.29 (1.11, 1.48) | 0.4 ng/ml (0.35, 0.45) | 6.99 |
| COV2-2130 | 293/ACE2 | HIV-1 | 1.6 ng/ml | (*38*) | 1.02 (0.77, 1.27) | 0.53 ng/ml (0.31, 0.9) | 5.39 | 0.96 (0.28, 1.64) | 0.62 ng/ml (0.29, 1.3) | 5.01 |
| P2C-1F11 | Huh7 | HIV-1 | 0.03 µg/ml | (*39*) | 0.85 (0.71, 1) | 0.02 µg/ml (0.01, 0.03) | 3.19 | 0.83 (0.67, 0.99) | 0.02 µg/ml (0.02, 0.03) | 2.99 |
| P2B-2F6 | Huh7 | HIV-1 | 0.05 µg/ml | (*39*) | 0.67 (0.58, 0.76) | 0.1 µg/ml (0.06, 0.16) | 2.02 | 0.64 (0.55, 0.73) | 0.06 µg/ml (0.05, 0.08) | 2.07 |
| P2C-1A3 | Huh7 | HIV-1 | 0.62 µg/ml | (*39*) | 0.56 (0.44, 0.67) | 0.44 µg/ml (0.25, 0.77) | 1.33 | 0.51 (0.34, 0.67) | 0.49 µg/ml (0.27, 0.9) | 1.2 |
| P2C-1C10 | Huh7 | HIV-1 | 2.62 µg/ml | (*39*) | 2.25 (1.64, 2.87) | 2.22 µg/ml (1.73, 2.86) | 3.73 | 2.3 (1.47, 3.13) | 2.26 µg/ml (1.93, 2.66) | 3.79 |
| P2B-2G4 | Huh7 | HIV-1 | 5.11 µg/ml | (*39*) | 0.8 (0.43, 1.17) | 10.57 µg/ml (5.95, 18.77) | 0.85 | 0.82 (0.35, 1.3) | 10.23 µg/ml (5.62, 18.62) | 0.88 |

| | | | | | | | | | | |
|---|---|---|---|---|---|---|---|---|---|---|
| P2A-1A8 | Huh7 | HIV-1 | 7.68 μg/ml | (*39*) | 0.85 (0.71, 1) | 5.5 μg/ml (3.97, 7.6) | 1.11 | 0.82 (0.65, 0.99) | 6.01 μg/ml (4.64, 7.8) | 1.04 |
| P2A-1A10 | Huh7 | HIV-1 | 8.57 μg/ml | (*39*) | 0.52 (0.31, 0.72) | 4.83 μg/ml (1.84, 12.67) | 0.76 | 0.56 (0.29, 0.84) | 6.29 μg/ml (2.8, 14.14) | 0.76 |
| BD-217 | Huh7 | VSV | 0.031 μg/ml | (*40*) | 1.14 (0.95, 1.33) | 0.05 μg/ml (0.04, 0.06) | 3.82 | 1.19 (0.98, 1.4) | 0.04 μg/ml (0.04, 0.05) | 3.99 |
| BD-218 | Huh7 | VSV | 0.011 μg/ml | (*40*) | 0.71 (0.59, 0.83) | 0.006 μg/ml (0.004, 0.01) | 2.99 | 0.68 (0.48, 0.89) | 0.009 μg/ml (0.005, 0.01) | 2.78 |
| BD-236 | Huh7 | VSV | 0.037 μg/ml | (*40*) | 0.83 (0.67, 1) | 0.03 μg/ml (0.02, 0.05) | 2.94 | 0.81 (0.7, 0.92) | 0.04 μg/ml (0.03, 0.05) | 2.78 |
| BD-361 | Huh7 | VSV | 0.02 μg/ml | (*40*) | 0.71 (0.62, 0.81) | 0.04 μg/ml (0.03, 0.06) | 2.42 | 0.74 (0.61, 0.87) | 0.04 μg/ml (0.03, 0.05) | 2.51 |
| BD-368[$] | Huh7 | VSV | 0.035 μg/ml | (*40*) | 0.72 (0.6, 0.84) | 0.04 μg/ml (0.02, 0.08) | 2.42 | 0.97 (0.79, 1.15) | 0.04 μg/ml (0.03, 0.05) | 3.3 |
| BD-368-2[$] | Huh7 | VSV | 0.0012 μg/ml | (*40*) | 1.16 (0.86, 1.46) | 0.002 μg/ml (0.001, 0.003) | 5.5 | 1.46 (1.33, 1.59) | 0.0018 μg/ml (0.0017, 0.0019) | 6.9 |
| BD-395 | Huh7 | VSV | 0.02 μg/ml | (*40*) | 0.96 (0.64, 1.28) | 0.02 μg/ml (0.01, 0.05) | 3.59 | 1.13 (0.79, 1.47) | 0.02 μg/ml (0.02, 0.03) | 4.09 |
| REGN10933 | Vero | VSV | 4.28 x $10^{-11}$ M | (*41*) | 1.06 (0.81, 1.31) | 6.17 x $10^{-11}$ M (4.11, 9.25) x $10^{-11}$ | 4.27 | 1.1 (0.86, 1.34) | 5.93 x $10^{-11}$ M (4.82, 7.29) x $10^{-11}$ | 4.45 |
| REGN10933 | Calu-3 | VSV | - | (*41*) | 1.0 (0.86, 1.14) | 1.58 x $10^{-11}$ M (1.11, 2.24) x $10^{-11}$ | 4.62 | 0.9 (0.62, 1.19) | 1.67 x $10^{-11}$ M (1.18, 2.36) x $10^{-11}$ | 4.16 |
| REGN10934 | Vero | VSV | 5.44 x $10^{-11}$ M | (*41*) | 1.46 (1.09, 1.84) | 9.62 x $10^{-11}$ M (6.45, 14.3) x $10^{-11}$ | 5.61 | 1.28 (0.98, 1.57) | 8.54 x $10^{-11}$ M (6.98, 10.5) x $10^{-11}$ | 4.96 |

| | | | | | | | | | | |
|---|---|---|---|---|---|---|---|---|---|---|
| REGN10934 | Calu-3 | VSV | - | (*41*) | 0.92 (0.64, 1.2) | $3.01 \times 10^{-11}$ M (1.85, 4.9) $\times 10^{-11}$ | 4.01 | 0.83 (0.45, 1.2) | $3.22 \times 10^{-11}$ M (1.9, 5.44) $\times 10^{-11}$ | 3.58 |
| REGN10954 | Vero | VSV | $9.22 \times 10^{-11}$ M | (*41*) | 1.45 (1.28, 1.63) | $3.07 \times 10^{-10}$ M (2.55, 3.7) $\times 10^{-10}$ | 4.85 | 1.41 (1.19, 1.63) | $3.2 \times 10^{-10}$ M (2.83, 3.62) $\times 10^{-10}$ | 4.69 |
| REGN10964[$] | Vero | VSV | $5.7 \times 10^{-11}$ M | (*41*) | 1.02 (0.67, 1.36) | $6.95 \times 10^{-11}$ M (2.62, 18.4) $\times 10^{-11}$ | 4.05 | 1.32 (1.15, 1.48) | $9.25 \times 10^{-11}$ M (8.32, 10.3) $\times 10^{-11}$ | 5.07 |
| REGN10977 | Vero | VSV | $5.15 \times 10^{-11}$ M | (*41*) | 1.11 (0.82, 1.41) | $7.74 \times 10^{-11}$ M (4.34, 13.8) $\times 10^{-11}$ | 4.38 | 1.1 (0.89, 1.3) | $1 \times 10^{-10}$ M (0.83, 1.21) $\times 10^{-11}$ | 4.19 |
| REGN10984[$] | Vero | VSV | $9.73 \times 10^{-11}$ M | (*41*) | 0.67 (0.35, 0.98) | $4.19 \times 10^{-11}$ M (1.07, 16.3) $\times 10^{-11}$ | 2.81 | 0.98 (0.57, 1.39) | $1.03 \times 10^{-10}$ M (0.67, 1.6) $\times 10^{-10}$ | 3.74 |
| REGN10986[$] | Vero | VSV | $9.91 \times 10^{-11}$ M | (*41*) | 0.81 (0.48, 1.14) | $8.09 \times 10^{-11}$ M (2.89, 22.7) $\times 10^{-11}$ | 3.18 | 1.21 (0.62, 1.8) | $1.82 \times 10^{-10}$ M (1.16, 2.87) $\times 10^{-10}$ | 4.31 |
| REGN10987[$] | Vero | VSV | $4.06 \times 10^{-11}$ M | (*41*) | 0.94 (0.74, 1.14) | $5.69 \times 10^{-11}$ M (3.03, 10.7) $\times 10^{-11}$ | 3.82 | 1.15 (1.04, 1.25) | $6.05 \times 10^{-11}$ M (5.54, 6.61) $\times 10^{-11}$ | 4.63 |
| REGN10987 | Calu-3 | VSV | - | (*41*) | 0.61 (0.55, 0.68) | $1.71 \times 10^{-11}$ M (1.06, 2.76) $\times 10^{-11}$ | 2.82 | 0.65 (0.56, 0.74) | $1.93 \times 10^{-11}$ M (1.57, 2.37) $\times 10^{-11}$ | 2.96 |
| REGN10989 | Vero | VSV | $7.23 \times 10^{-12}$ M | (*41*) | 0.84 (0.5, 1.19) | $7.60 \times 10^{-12}$ M (3.52, 16.4) $\times 10^{-12}$ | 4.17 | 0.94 (0.6, 1.29) | $1 \times 10^{-11}$ M (0.66, 1.52) $\times 10^{-11}$ | 4.56 |
| REGN10989 | Calu-3 | VSV | - | (*41*) | 1.26 (1.09, 1.43) | $7.85 \times 10^{-12}$ M (6.22, 9.91) $\times 10^{-12}$ | 6.21 | 1.19 (0.91, 1.48) | $7.85 \times 10^{-12}$ M (6.52, 9.45) $\times 10^{-12}$ | 5.89 |
| CC6.29[$*] | HeLa/ACE2 | MLV | 0.002 µg/ml | (*42*) | 0.86 (0, 1.77) | 0.003 µg/ml (0.0004, 0.02) | 3.91 | 0.66 (0, 1.41) | 0.002 µg/ml (0.0004, 0.01) | 3.09 |
| CC6.30[$] | HeLa/ACE2 | MLV | 0.0013 µg/ml | (*42*) | 0.66 (0.33, 0.98) | 0.001 µg/ml (0.0001, 0.002) | 3.47 | 0.55 (0.21, 0.88) | 0.001 µg/ml (0.0003, 0.003) | 2.73 |

| | | | | | | | | | | |
|---|---|---|---|---|---|---|---|---|---|---|
| CC6.31* | HeLa/ACE2 | MLV | 0.059 µg/ml | (*42*) | 0.41 (0, 1.45) | 0.55 µg/ml (0.005, 64.13) | 0.97 | 0.35 (0, 1.38) | 0.71 µg/ml (0.0048, 105.98) | 0.82 |
| CC6.33 | HeLa/ACE2 | MLV | 0.039 µg/ml | (*42*) | 0.6 (0.37, 0.83) | 0.02 µg/ml (0.01, 0.06) | 2.27 | 0.66 (0.4, 0.93) | 0.03 µg/ml (0.02, 0.06) | 2.31 |
| CC12.1* | HeLa/ACE2 | MLV | 0.019 µg/ml | (*42*) | 1.14 (0, 2.85) | 0.02 µg/ml (0.003, 0.14) | 4.23 | 1.11 (0, 3.49) | 0.02 µg/ml (0.002, 0.12) | 4.18 |
| CC12.3 | HeLa/ACE2 | MLV | 0.018 µg/ml | (*42*) | 1.02 (0.6, 1.43) | 0.02 µg/ml (0.01, 0.05) | 3.76 | 0.88 (0.64, 1.12) | 0.02 µg/ml (0.01, 0.03) | 3.28 |
| CC12.4 | HeLa/ACE2 | MLV | 0.11 µg/ml | (*42*) | 0.95 (0.62, 1.28) | 0.08 µg/ml (0.04, 0.18) | 2.95 | 0.86 (0.49, 1.23) | 0.11 µg/ml (0.06, 0.19) | 2.55 |
| CC12.14 | HeLa/ACE2 | MLV | 0.023 µg/ml | (*42*) | 0.79 (0.67, 0.91) | 0.03 µg/ml (0.02, 0.04) | 2.78 | 0.81 (0.7, 0.91) | 0.04 µg/ml (0.03, 0.05) | 2.76 |
| ADI-55688$ | HeLa/ACE2 | MLV | - | (*35*) | 0.45 (0.15, 0.74) | 0.82 nM (0.07, 10.24) | 1.32 | 0.81 (0.38, 1.23) | 1.26 nM (0.62, 2.55) | 2.2 |
| ADI-55688 | Vero | VSV | - | (*35*) | 0.82 (0.6, 1.04) | 0.37 nM (0.16, 0.84) | 2.67 | 0.88 (0.45, 1.3) | 0.46 nM (0.25, 0.83) | 2.77 |
| ADI-55689 | HeLa/ACE2 | MLV | - | (*35*) | 0.72 (0.43, 1.01) | 0.85 nM (0.28, 2.59) | 2.08 | 0.79 nM (0.35, 1.22) | 1.63 nM (0.74, 3.58) | 2.06 |
| ADI-55689 | Vero | VSV | - | (*35*) | 0.77 (0.47, 1.07) | 0.16 nM (0.08, 0.32) | 2.79 | 0.76 (0.27, 1.24) | 0.14 nM (0.07, 0.31) | 2.78 |
| ADI-55690$ | HeLa/ACE2 | MLV | - | (*35*) | 0.44 (0.12, 0.75) | 0.64 nM (0.02, 20.49) | 1.34 | 0.79 (0.1, 1.48) | 4.48 nM (1.53, 13.08) | 1.73 |
| ADI-55690 | Vero | VSV | - | (*35*) | 0.73 (0.49, 0.96) | 1.27 nM (0.48, 3.39) | 1.98 | 0.65 (0.24, 1.06) | 1.56 nM (0.58, 4.19) | 1.72 |

| | | | | | | | | | | |
|---|---|---|---|---|---|---|---|---|---|---|
| ADI-55951$ | HeLa/ACE2 | MLV | - | (*35*) | 0.24<br>(0.01, 0.48) | 17.21 nM<br>(0.01, 33973.11) | 0.54 | 0.64<br>(0.13, 1.16) | 10.46 nM<br>(3.35, 32.72) | 1.19 |
| ADI-55951$ | Vero | VSV | - | (*35*) | 0.87<br>(0.37, 1.36) | 2.33 nM<br>(0.57, 9.49) | 2.13 | 1.42<br>(0.56, 2.29) | 5.03 nM<br>(3.1, 8.15) | 3.02 |
| ADI-55993$ | HeLa/ACE2 | MLV | - | (*35*) | 0.38<br>(0.15, 0.61) | 0.31 nM<br>(0.02, 6.48) | 1.28 | 0.71<br>(0.28, 1.15) | 0.68 nM<br>(0.27, 1.72) | 2.14 |
| ADI-55993 | Vero | VSV | - | (*35*) | 1.04<br>(0.55, 1.53) | 0.45 nM<br>(0.19, 1.08) | 3.31 | 0.99<br>(0.43, 1.55) | 0.45 nM<br>(0.26, 0.79) | 3.15 |
| ADI-56000$ | HeLa/ACE2 | MLV | - | (*35*) | 0.43<br>(0.21, 0.65) | 0.14 nM<br>(0.01, 1.64) | 1.6 | 0.92<br>(0.24, 1.6) | 0.68 nM<br>(0.29, 1.61) | 2.74 |
| ADI-56000 | Vero | VSV | - | (*35*) | 1.37<br>(0.76, 1.97) | 0.75 nM<br>(0.33, 1.71) | 4.04 | 1.45<br>(0.52, 2.39) | 0.61 nM<br>(0.37, 1.02) | 4.41 |
| ADI-56010$ | HeLa/ACE2 | MLV | - | (*35*) | 0.38<br>(0.15, 0.61) | 0.25 nM<br>(0.01, 4.9) | 1.33 | 0.73<br>(0.28, 1.17) | 1.55 nM<br>(0.6, 3.97) | 1.92 |
| ADI-56010 | Vero | VSV | - | (*35*) | 1.15<br>(1.05, 1.25) | 1.79 nM<br>(1.46, 2.19) | 2.95 | 1.13<br>(1.01, 1.24) | 1.96 nM<br>(1.78, 2.17) | 2.85 |
| ADI-56046 | HeLa/ACE2 | MLV | - | (*35*) | 0.71<br>(0.38, 1.05) | 0.86 nM<br>(0.25, 2.95) | 2.07 | 0.64<br>(0.24, 1.04) | 1 nM<br>(0.39, 2.56) | 1.82 |
| ADI-56046 | Vero | VSV | - | (*35*) | 1.54<br>(0.99, 2.09) | 0.44 nM<br>(0.25, 0.77) | 4.9 | 1.41<br>(0.37, 2.44) | 0.33 nM<br>(0.19, 0.6) | 4.64 |
| COV21 IgG$ | 293T/ACE2 | HIV-1 | 62.3 nM | (*43*) | 1.44<br>(0.6, 2.28) | 45.23 nM<br>(17.3, 118.25) | 1.69 | 1.22<br>(0.67, 1.78) | 55.8 nM<br>(37.04, 84.06) | 1.34 |
| COV57 IgG* | 293T/ACE2 | HIV-1 | 121.1 nM | (*43*) | 1.25<br>(0, 4.85) | 111.25 nM<br>(3.66, 3384.5) | 1.01 | 1.12<br>(0, 5.54) | 128.92 nM<br>(4.1, 4059.6) | 0.86 |

| | | | | | | | | | | |
|---|---|---|---|---|---|---|---|---|---|---|
| S309$ | VeroE6 or DBT/ACE2 | MLV | - | (*44*) | 0.91 (0.47, 1.34) | 1.5 ng/ml (0.57, 3.94) | 4.38 | 1.18 (0.71, 1.66) | 1.08 ng/ml (0.75, 1.55) | 5.89 |
| S309$ | VeroE6 or DBT/ACE2 | MLV | - | (*44*) | 0.85 (0.55, 1.14) | 1.05 ng/ml (0.44, 2.51) | 4.22 | 1.07 (0.64, 1.49) | 0.5 ng/ml (0.33, 0.76) | 5.65 |
| Ty1-Fc | 293T/ACE2 | - | 0.012 μg/ml | (*45*) | 0.53 (0.46, 0.61) | 0.02 μg/ml (0.01, 0.03) | 1.96 | 0.57 (0.43, 0.72) | 0.02 μg/ml (0.01, 0.03) | 2.13 |
| Ty1 | 293T/ACE2 | - | 0.77 μg/ml | (*45*) | 0.84 (0.8, 0.88) | 0.89 μg/ml (0.79, 1) | 1.74 | 0.85 (0.77, 0.93) | 0.83 (0.74, 0.94) | 1.78 |

$NAbs ignored because $IIP_{100}$ values estimated using Eqs. [1] and [2] deviated by 20% or more
*For 8 NAbs, the lower limit of 95% confidence interval was negative and was truncated to 0
@We assumed the molecular weight of NAbs to be 150 kDa for unit conversions
### $IC_{50}$ values were digitized from Ref. (*6*)

**Table S2: Viral dynamics model parameters and their values.**

| Parameter | Description | Value (Range) | Ref. |
|---|---|---|---|
| $b$ | Infection rate constant of cells | 6 x $10^{-8}$ virions$^{-1}$ ml d$^{-1}$<br>(2.4 - 15.1) x $10^{-8}$ | (*24*) |
| $r_X$ | Refractory state conversion rate constant | 4 d$^{-1}$<br>(3 - 5) | (*51*) |
| $d$ | Infected cell death<br>rate constant | 0.6 d$^{-1}$<br>(0.1 - 1.1) | (*25*) |
| $p$ | Virion production rate per infected cell | 390 virions ml$^{-1}$ d$^{-1}$<br>(290 - 490) | (*24*) |
| $c$ | Virion clearance rate constant | 20 d$^{-1}$<br>(15 - 20) | (*24*) |
| $s_X$ | Maximal innate immunity activation rate constant | 1 d$^{-1}$<br>(0.5 - 1.5) | (*51*) |
| $f_X$ | Infected cell numbers at which innate immunity activation is half-maximal | 100 cells<br>(0.01 - 200) | (*51*) |
| $d_X$ | Innate immunity decay rate constant | 0.2 d$^{-1}$<br>(0.15 - 0.25) | (*51*) |
| $T(0)$ | Initial target cells | 3 x $10^7$ cells | Fixed |
| $I(0)$ | Initial infected cells | 1 cell | (*24*) |
| $V(0)$ | Initial viral load | $\frac{pI(0)}{c}$ | (*24*) |
| $X(0)$ | Initial innate immune reponse<br>(normalised value) | 0 | Fixed |

[#]The ranges of parameter values used in simulations to account for inter-patient variations are indicated in parentheses

**Table S3. Summary of clinical trials.**

| Vaccine | $NT_{50}$ (95% CI) | Efficacy (95% CI) | Refs. |
|---|---|---|---|
| BNT162b2* (1 dose)** | 14 (10 – 20) | 49 (41 – 57) | (*55*, *56*) |
| BNT162b2* (2 doses)*** | 361 (235 – 550) | 94 (87 – 97) | (*55*, *56*) |
| mRNA-1273& (2 doses) | 356.2 (262.6 – 483.1) | 95.6 (90.6 – 97.9) | (*1*, *57*) |
| ChAdOx1 nCoV-19$ (2 doses) | 239 (210 – 275) | 80 (65.2 – 88.5) | *(58)* |
| Gam-COVID-Vac (Sputnik V)# (2 doses) | 44.5 (31.8 – 62.2) | 91.6 (85.6 – 95.2) | (*4*) |

*$NT_{50}$ measurements were from individuals aged between 18 and 55 years (*56*). Efficacy measurement included individuals aged between 16 and 39 years

**$NT_{50}$ was measured on day 21 after 1st dose and efficacy was measured between days 14 and 21

***$NT_{50}$ was measured on day 7 after 2nd dose (day 28 after 1st dose) and efficacy was measured 7 days after 2nd dose to end of the study

#$NT_{50}$ was measured on day 42 after first dose and efficacy was measured from day 21 after 1st dose (the day of 2nd dose)

&$NT_{50}$ measurements were from individuals aged between 18 and 55 years. Efficacy measurement included individuals aged between 18 and 65 years. $NT_{50}$ values were digitized using Engauge 12.1.

$The interval between doses was ≥12 weeks. $NT_{50}$ values were digitized using Engauge 12.1
